# Systematic Review and Meta-Analysis Protocol: Beta adrenoreceptor drugs and risk of Parkinson’s disease

**DOI:** 10.1101/2021.05.19.21257436

**Authors:** Ambrish Singh, Salman Hussain, Sreelatha Akkala, Jitka Klugarová, Andrea Pokorná, Miloslav Klugar, Benny Antony

## Abstract

Parkinson’s disease (PD) is a progressive nervous system disorder characterised by the loss of dopaminergic neurons leading to motor and non-motor symptoms. Accumulation of α-synuclein protein (SNCA) in the form of Lewy bodies has been observed in dopaminergic neurons of PD patients. Potential relationships between β-adrenergic drugs (agonists and antagonist) and SNCA synthesis in PD have been recently suggested. This study aims to systematically review the evidence from various epidemiological studies that analysed the association between beta-adrenoceptors (agonists and antagonists) and the risk of PD. Biomedical databases such as PubMed and Embase will be searched to identify the individual studies that reported the relationship between beta-adrenoceptors and the risk of PD. JBI critical appraisal tool scale will be used to assess the quality of included studies. The primary outcome will be to compute the pooled risk of PD among beta-agonist and antagonist users. Furthermore, we will consider the pooled risk of PD based on study design, types of beta-agonist or antagonist exposure under secondary outcomes. RevMan 5, STATA 16, and ProMeta 3.0 will be used to conduct the statistical analysis.

## Background and rationale

Parkinson’s disease (PD) is a neurodegenerative disorder that leads to progressive deterioration of motor function due to loss of dopaminergic neurons. PD is characterized by the cardinal features of rest tremor, bradykinesia, rigidity, postural instability, and a variety of other motor and non-motor symptoms.^1,2^ Globally, an estimated 9.4 million people live with PD. With an aging population, the prevalence and incidence of PD are expected to increase by >30% by 2030, contributing to a significant humanistic and economic burden globally.^3,4^

Although the etiology of PD remains unclear, a variety of genetic and environmental factors have been identified as risk (aging, family history, pesticide exposure) or protective (tobacco) factors. The α-synuclein gene (SNCA) is a presynaptic neuronal protein that is linked genetically to PD and suggested to play an important role in the disease process. In addition, SNCA is present in Lewy bodies, the neuropathological hallmark of PD, which have been linked to neurotoxic pathways leading to neurodegeneration.^5^

Current neuroprotective therapies target the spread, production, aggregation, and degradation of SNCA.^5^ Recent *in vitro* and animal model studies have found associations of beta-adrenoreceptor with neuronal SNCA expression and demonstrated a regulatory action of the SNCA through beta-adrenoceptor activation.^1,6^ This evidence has been strengthened by recent observational studies that investigated the association between beta-adrenoceptors (agonists and antagonists) exposure and PD onset. ^7-9^ The study results demonstrated that the chronic use of the beta-adrenoceptor antagonist (propranolol) was associated with an increased risk of PD, while the chronic use of the beta-adrenoceptor agonists (salbutamol) was associated with a decreased risk.^8^ Several studies reported the association between beta-adrenoceptors and the risk of PD; however, the findings of these studies were conflicting. In addition, no systematic review has assessed the effect of beta-adrenoceptors (agonists and antagonists separately) use on the risk of PD.

### Objectives

We aim to systematically review the evidence from epidemiological studies to understand the association and meta-analyse the pooled relative risk (RR) of PD with beta-adrenoceptors (agonist and antagonist separately) exposure.

## Methodology

### Eligibility criteria

We will select the eligible epidemiological studies following the specific criteria of PICOS items: Participants, Interventions, Comparators, Outcomes, and Study design. The present systematic review and meta-analysis will be conducted and reported according to the recommendations of the preferred systematic review and meta-analysis (PRISMA) and Meta-analysis of Observational Studies in Epidemiology (MOOSE) reporting guideline.^10,11^

### Characteristics of participants (P)

We will include patients of any age and receiving beta-agonist or antagonist and have confirmed diagnosis of PD.

### Characteristics of intervention/exposures (I)

This study will focus on patients who were on beta agonist (salbutamol; terbutaline; formoterol; salmeterol; indacterol; vilanterol; olodaterol) and beta-antagonist (Bisoprolol; Nebivolol; Atenolol; Acebutolol; Sotalol; Propranolol; Celiprolol; Metoprolol; Pindolol; Betaxolol; Nadolol; Carvedilol; Tertatolol; Labetalol; Timolol)

### Characteristics of comparators (C)

We will include all the studies assessing the risk of PD in beta-agonist and/or antagonist users compared to non-users or any other active drug users.

### Outcomes of interest (O)

Our primary outcome is to compute the risk of PD among beta-agonist and/or antagonist users. Studies reporting the risk ratio, odds ratio (OR) and hazard ratio (HR) or the absolute numbers to estimate these measures will be of interest. Studies that qualify all other inclusion criteria and assessed any of these outcomes but did not report the data or reported the data in a format not suitable for quantitative synthesis, we will include such studies in the review and present relevant information in the narrative form.

### Characteristics of study design (S)

Epidemiological analytical studies such as retrospective studies, prospective studies, cohort studies, case-control studies will be included in this meta-analysis. Studies reporting reviews, case series, case reports, genetic studies, animal studies, commentary, editorial, or study protocol will be excluded.

## Information sources and search procedure

We will implement the search procedure consistently with the following criteria.

### Electronic source and search strategy

For this systematic review and meta-analysis, a three-step search strategy will be utilized to locate both published and unpublished studies. An initial limited search will be undertaken in MEDLINE (Ovid) and Embase (Ovid), using keywords and index terms related to Parkinson’s disease, β2 adrenergic receptor agonists, β2 adrenergic receptor antagonists, risk factors or incidence. An analysis of the text words contained in the title and abstract and the index terms used to describe the articles will follow. A second search using all identified keywords and index terms will be conducted across all included databases. Thirdly, the reference lists of all studies that will meet the inclusion criteria will be checked for additional records.

The databases to be searched include MEDLINE (Ovid), Embase (Ovid), Scopus, Web of Science Core Collection, Emcare (Ovid) and CINAHL (EBSCO). Sources of grey literature will be ProQuest Dissertations & Theses Global and clinical trials registers ClinicalTrials.gov and WHO International Clinical Trials Registry Platform (ICTRP). No language restriction will be applied during the search; however, only the studies published in English language will be included.

### Hand-searching

Abstract booklet from conference proceedings and poster sessions, from last two years, will be hand searched using the online sources of major international association involved in PD research: American Academy of Neurology (AAN), International Parkinson and Movement Disorder Society (MDS), and World Federation of Neurology (WFN). Furthermore, the reference list of relevant studies and pertinent review articles will be searched additionally to identify other studies meeting selection criteria.

### Study selection

An Inclusion/Exclusion Form was adapted and will be used for screening (Appendix 1). Two reviewers independently judged the study against the inclusion and exclusion criteria based on title and abstract screening in the initial phase and full-text screening in the later phase. Each potential discrepancy will be discussed and solved through consensus with other authors and independent expert consultation.

### Assessment of risk of bias

The methodological quality of the selected studies will be evaluated using the Jonna Briggs Institute (JBI) critical appraisal tool.^12^ A Risk-of-Bias form was adapted and will be used for data extraction (Appendix 2). Each article will be evaluated independently by two researchers using the JBI tool. Depending on the response, each domain of the tool will be classified as high, low or unclear risk of bias.

### Data extraction

A standard data extraction form will be adapted and used for data extraction (Appendix 3). Two reviewers will extract data independently from the included studies for the following information: study design, characteristics of the population (age, sex, and male percentage), sample size, intervention details, duration of follow-up, outcome measurements, The missing data in the literature will be obtained by emailing the corresponding author; otherwise, it will be estimated by the appropriate method according to the Cochrane Handbook 5.1.0.^13^

## Statistical analysis

The primary outcome of this study is to compute the pooled RR of PD among beta-agonist and/or antagonist users, separately. A generic inverse variance method will be applied to compute the overall RR of PD among beta-agonist and antagonist users. Considering the incidence of PD to be rare or low, the risk ratio, OR, and HR will be considered an equivalent measure of risk; therefore, we will use RR representing all of these measures for simplicity.^14^ Heterogeneity among the included studies will be assessed using Cochrane chisquared and I^2^ tests. Cochrane chi-squared value (p< 0.10) and I^2^ value ≥50% may represent considerable heterogeneity.^15^ Based on the heterogeneity assessment, a random-effect or fixed-effect model will be chosen. If considerable heterogeneity observed, then the analysis will be performed using a random-effect model. Subgroup analysis will be performed based on study design, types of beta-agonist or antagonist exposure, and duration of exposure. Sensitivity analysis will be carried out by omitting a single study one by one (leave-one-out method) from the pooled analysis, and the risk of bias on the outcome will also be explored using the sensitivity analysis accordingly to the high, medium, or low risk.^16^ Publication bias will be detected based on the visual inspection of the funnel plot, and the trim-and-fill method will be used to estimate the effect of publication bias (if any). Meta-analysis will be performed using Review Manager version 5.4.1. (Copenhagen: The Nordic Cochrane Centre, The Cochrane Collaboration, 2014), STATA version 16 (STATA Corp., Texas, USA), and ProMeta Version 3.0 ([Computer software]. Cesena, Italy: Internovi).

### Certainty of Evidence

The potential for publication bias will be assessed using a funnel plot construction if there are 10 or more studies. The y-axis will be the study population size, and the x-axis will be the measure of effect for that property. If a meta-analysis is conducted, the potential for small-study effects will be evaluated by comparing the fixed-effects model to the random-effects model. If the effects are similar, then small study effects are unlikely. If the random-effects model is more beneficial, then we will consider the potential for the results to be different in the smaller studies. As per COSMIN guidelines for systematic reviews, the quality of evidence will be assessed with the Grading of Recommendations Assessment, Development and Evaluation (GRADE) using the five domains: risk of bias, consistency, directness, precision, and publication bias.^17^ Reviewers assessing the certainty of evidence will refer to the COSMIN handbook to ensure proper utilization of the tool for systematic reviews of exposures. The overall certainty of evidence will be considered high (a lot of confidence that the true effect is similar to the estimated effect), moderate (the true effect is probably close to the estimated effect), low (true effect might be markedly different from the estimated effect), or very low (the true effect is probably markedly different from the estimated effect) for each outcome accordingly.

## Data Availability

Not applicable

## Appendix 1

### Inclusion/Exclusion Form for Primary Studies

Study ID:

Reviewer:

Date:

**Identification Details**

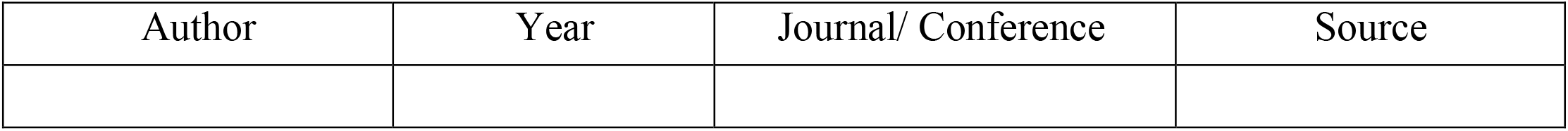

On Endnote database ..................................Yes / No Full

Text availability ..................................Yes / No

**Study Eligibility**

Study design is one of the following:

Case-control/Cohort..................................................................................Yes/ No

The study concerns PD............................................................................... Yes/ No

The study concerns Beta-agonist or antagonist use.................................................... Yes/ No

The study is a human study, not animal/laboratory experiment......................................... Yes/ No

#### Please Tick Only One Box Below

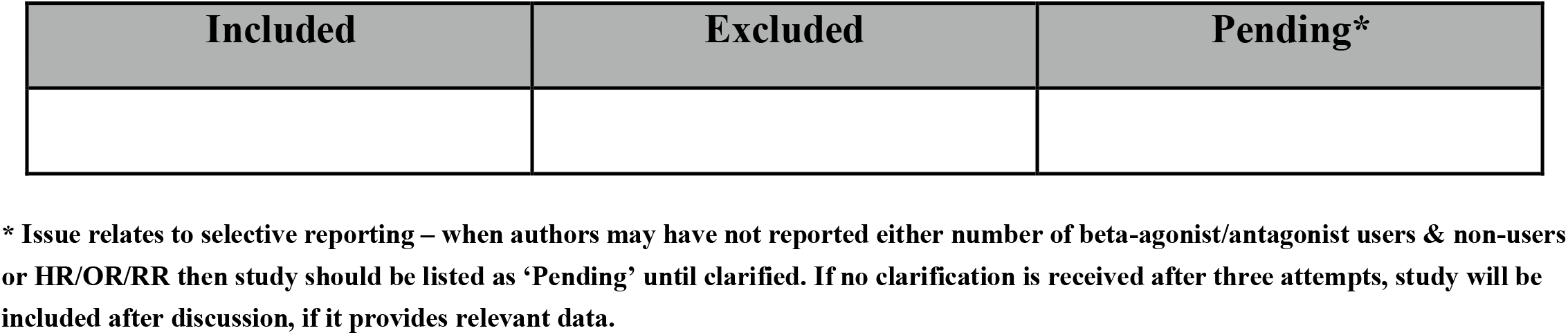

#### References to Other studies

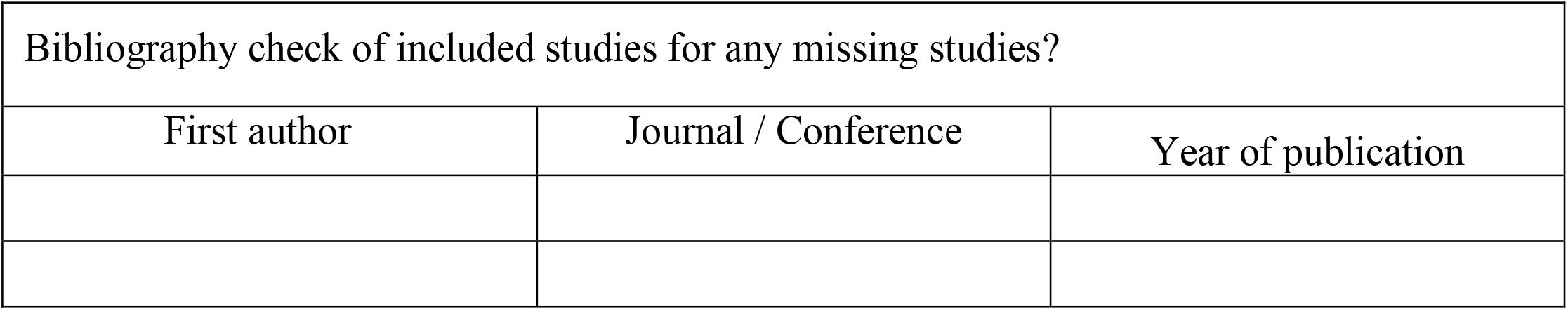

| Bibliography check of included studies for any missing studies? |  |  |
| --- | --- | --- |
| First author | Journal / Conference | Year of publication |

## Appendix 2

**Risk-of-Bias Form**

### JBI CRITICAL APPRAISAL CHECKLIST FOR COHORT STUDIES

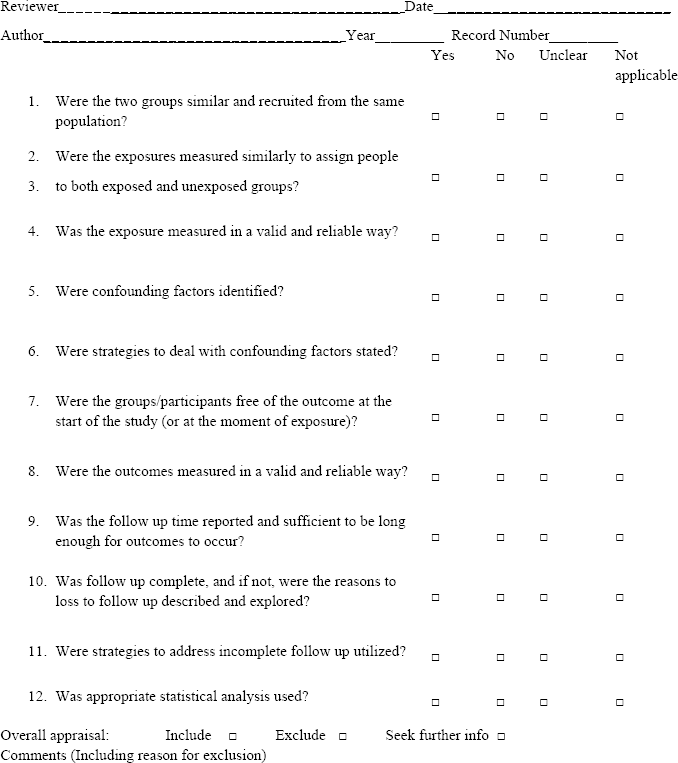

### JBI CRITICAL APPRAISAL CHECKLIST FOR CASE CONTROL STUDIES

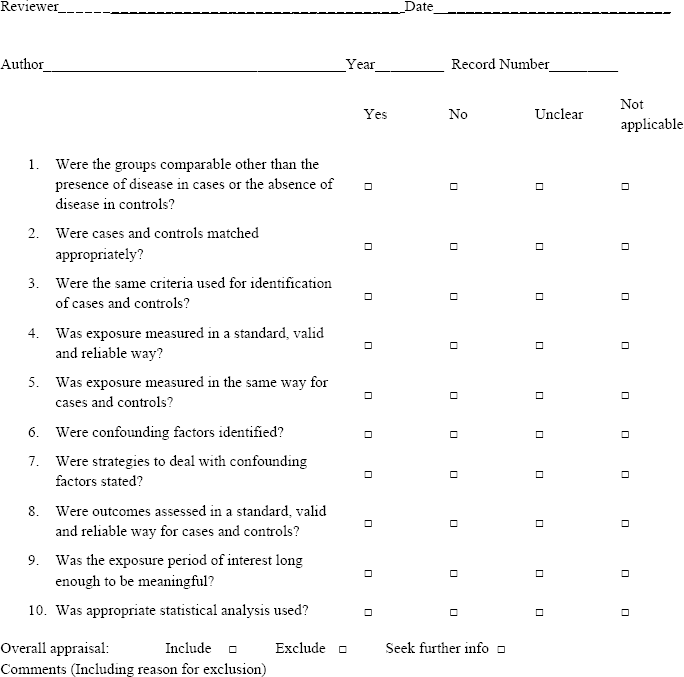

## Appendix 3

### Data Extraction Form

#### Study and Participants characteristics

| Study characteristics |  |
| --- | --- |
|  | Further details |
| Study author; year |  |
| Country / Countries |  |
| Study design |  |
| Database used |  |
| How was participant eligibility defined? |  |
| What was the sample size? |  |
| How the use of beta-agonist and antagonist was assessed? |  |
| How PD was ascertained? |  |
| Duration of follow-up |  |
| Other |  |
| Participant characteristics |  |
|  | Further details |
| Number of participants (cases versus controls/Cohort size) |  |
| Age (mean, median, range, etc) |  |
| BMI (mean, median, range, etc) |  |
| Male participants (numbers / %, etc) |  |
| Smoker/non-smoker |  |
| Other |  |

| Measures relevant to the review |  |  |  |  |
| --- | --- | --- | --- | --- |
| HR |  | YES/ NO |  |  |
| OR |  | YES/ NO |  |  |
| RR |  | YES/ NO |  |  |
| Others |  |  |  |  |
| For dichotomous data |  |  |  |  |
| HR/OR/ RR<br>(95% CI, p value) | Exposure group (n) |  | Non-exposure group (n) |  |
|  | N = number of participants; n=not number of events |  | N = number of participants; n=not number of events |  |
|  | N | n | N | n |
| Other Information (eg. Adjustment for confounding factors): |  |  |  |  |
| Other relevant data |  |  |  |  |

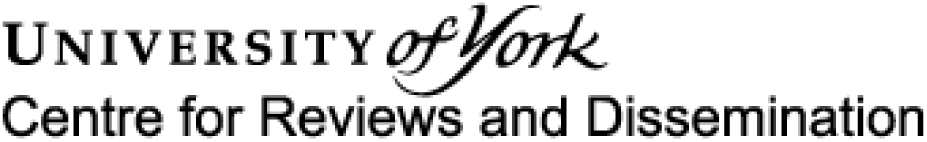

### Systematic review

*Fields that have an* ***asterisk (*)*** *next to them means that they* ***must be answered. Word limits*** *are provided for each section. You will be unable to submit the form if the word limits are exceeded for any section*.

*Registrant means the person filling out the form*.

#### 1. * Review title

Give the title of the review in English

Beta adrenoreceptor drugs and risk of Parkinson’s disease: A protocol for systematic review and meta-analysis

#### 2. Original language title

For reviews in languages other than English, give the title in the original language. This will be displayed with the English language title.

#### 3. * Anticipated or actual start date

Give the date the systematic review started or is expected to start. 03/11/2020

#### 4. * Anticipated completion date

Give the date by which the review is expected to be completed. 30/07/2021

#### 5. * Stage of review at time of this submission

Tick the boxes to show which review tasks have been started and which have been completed. Update this field each time any amendments are made to a published record.

**Reviews that have started data extraction (at the time of initial submission) are not eligible for inclusion in PROSPERO**. If there is later evidence that incorrect status and/or completion date has been supplied, the published PROSPERO record will be marked as retracted.

This field uses answers to initial screening questions. It cannot be edited until after registration.

The review has not yet started: No

| Review stage | Started | Completed |
| --- | --- | --- |
| Preliminary searches | Yes | No |
| Piloting of the study selection process | Yes | No |
| Formal screening of search results against eligibility criteria | Yes | No |
| Data extraction | No | No |
| Risk of bias (quality) assessment | Yes | No |
| Data analysis | Yes | No |

Provide any other relevant information about the stage of the review here.

#### 6. * Named contact

The named contact is the guarantor for the accuracy of the information in the register record. This may be any member of the review team.

Ambrish Singh

Email salutation (e.g. “Dr Smith” or “Joanne”) for correspondence:

Singh

#### 7. * Named contact email

Give the electronic email address of the named contact.

#### 8. Named contact address

Give the full institutional/organisational postal address for the named contact.

Menzies Institute for Medical Research, University of Tasmania, Hobart, Tasmania, Australia

#### 9. Named contact phone number

Give the telephone number for the named contact, including international dialling code.

+61 0469701341

#### 10. * Organisational affiliation of the review

Full title of the organisational affiliations for this review and website address if available. This field may be completed as ‘None’ if the review is not affiliated to any organisation.

Menzies Institute for Medical Research, University of Tasmania

Organisation web address:

#### 11. * Review team members and their organisational affiliations

Give the personal details and the organisational affiliations of each member of the review team. Affiliation refers to groups or organisations to which review team members belong. **NOTE: email and country now MUST be entered for each person, unless you are amending a published record**.

Mr Ambrish Singh. Menzies Institute for Medical Research, University of Tasmania

Dr Salman Hussain. Cochrane Czech Republic, Czech National Centre for Evidence-Based Healthcare and Knowledge Translation,(Czech EBHC: JBI Centre of Excellence, Masaryk University GRADE Centre), Institute of Biostatistics and Analyses, Faculty of Medicine, Masaryk University, Brno, Czech Republic Ms Sreelatha Akkala. Independent Researcher, Banglore, India

Dr Jitka Klugarová. Czech National Centre for Evidence-Based Healthcare and Knowledge Translation,(Cochrane Czech Republic, Czech EBHC: JBI Centre of Excellence, Masaryk University GRADE Centre), Institute of Biostatistics and Analyses, Faculty of Medicine, Masaryk University, Brno, Czech Republic

Dr Andrea Andrea Pokorná. Department of Nursing and Midwifery, Faculty of Medicine, Masaryk University, Brno, Czech Republic

Dr Miloslav Klugar. Cochrane Czech Republic, Czech National Centre for Evidence-Based Healthcare and Knowledge Translation,(Czech EBHC: JBI Centre of Excellence, Masaryk University GRADE Centre), Institute of Biostatistics and Analyses, Faculty of Medicine, Masaryk University, Brno, Czech Republic

Dr Benny Antony. Menzies Institute for Medical Research, University of Tasmania, Hobart, Tasmania, Australia

#### 12. * Funding sources/sponsors

Details of the individuals, organizations, groups, companies or other legal entities who have funded or sponsored the review.

No funding

##### Grant number(s)

State the funder, grant or award number and the date of award

#### 13. * Conflicts of interest

List actual or perceived conflicts of interest (financial or academic).

None

#### 14. Collaborators

Give the name and affiliation of any individuals or organisations who are working on the review but who are not listed as review team members. **NOTE: email and country must be completed for each person, unless you are amending a published record**.

#### 15. * Review question

State the review question(s) clearly and precisely. It may be appropriate to break very broad questions down into a series of related more specific questions. Questions may be framed or refined using PI(E)COS or similar where relevant.

Do beta-adrenoceptor agonists use associated with the risk of Parkinson’s disease (PD)?

Do beta-adrenoceptor antagonists use associated with the risk of PD?

Is there any association between the duration of beta-adrenoceptors (agonists and antagonists separately) exposure and the risk of PD?

What is the association between beta-adrenoceptors (agonists and antagonists separately) use and the risk of PD based on the type of beta-agonist or antagonist exposure?

#### 16. * Searches

State the sources that will be searched (e.g. Medline). Give the search dates, and any restrictions (e.g. language or publication date). Do NOT enter the full search strategy (it may be provided as a link or attachment below.)

A three-step search strategy will be utilized to locate both published and unpublished studies. An initial limited search will be undertaken in MEDLINE (Ovid) and Embase (Ovid), using keywords and index terms related to Parkinson’s disease, ?2 adrenergic receptor agonists, ?2 adrenergic receptor antagonists, risk factors or incidence. An analysis of the text words contained in the title and abstract and the index terms used to describe the articles will follow. A second search using all identified keywords and index terms will be conducted across all included databases. Thirdly, the reference lists of all studies that will meet the inclusion criteria will be checked=“checked” value=“1” for additional records.

Abstract booklet from conference proceedings and poster sessions, from last two years, will be hand searched using the online sources of major international association involved in PD research: American Academy of Neurology (AAN), International Parkinson and Movement Disorder Society (MDS), and World Federation of Neurology (WFN).

#### 17. URL to search strategy

Upload a file with your search strategy, or an example of a search strategy for a specific database, (including the keywords) in pdf or word format. In doing so you are consenting to the file being made publicly accessible. Or provide a URL or link to the strategy. Do NOT provide links to your search **results**.

Alternatively, upload your search strategy to CRD in pdf format. Please note that by doing so you are consenting to the file being made publicly accessible.

Do not make this file publicly available until the review is complete

#### 18. * Condition or domain being studied

Give a short description of the disease, condition or healthcare domain being studied in your systematic review.

Parkinson’s disease is a neurodegenerative disorder that leads to progressive deterioration of motor function due to loss of dopaminergic neurons. PD is characterized by the cardinal features of rest tremor, bradykinesia, rigidity, postural instability, and a variety of other motor and non-motor symptoms. Globally, an estimated 9.4 million people live with PD. With an aging population, the prevalence and incidence of PD are expected to increase by 30% by 2030, contributing to a significant humanistic and economic burden globally.

#### 19. * Participants/population

Specify the participants or populations being studied in the review. The preferred format includes details of both inclusion and exclusion criteria.

Patients of any age and receiving beta-agonist or antagonist and have confirmed diagnosis of PD.

#### 20. * Intervention(s), exposure(s)

Give full and clear descriptions or definitions of the interventions or the exposures to be reviewed. The preferred format includes details of both inclusion and exclusion criteria.

Patients who were on beta agonist (such as: salbutamol; terbutaline; formoterol; salmeterol; indacterol; vilanterol; olodaterol) and beta-antagonist (such as: bisoprolol; nebivolol; atenolol; acebutolol; sotalol; propranolol; celiprolol; metoprolol; pindolol; betaxolol; nadolol; carvedilol; tertatolol; labetalol; timolol)

#### 21. * Comparator(s)/control

Where relevant, give details of the alternatives against which the intervention/exposure will be compared (e.g. another intervention or a non-exposed control group). The preferred format includes details of both inclusion and exclusion criteria.

All the studies assessing the risk of PD in beta-agonist and/or antagonist users compared to non-users or any other active drug users.

#### 22. * Types of study to be included

Give details of the study designs (e.g. RCT) that are eligible for inclusion in the review. The preferred format includes both inclusion and exclusion criteria. If there are no restrictions on the types of study, this should be stated.

Epidemiological studies such as retrospective studies, prospective studies, cohort studies, case-control studies will be included in this meta-analysis.

Studies reporting reviews, case series, case reports, genetic studies, animal studies, commentary, editorial, or study protocol will be excluded.

#### 23. Context

Give summary details of the setting or other relevant characteristics, which help define the inclusion or exclusion criteria.

They assess the association between beta-adrenoceptors (agonists and antagonists separately) use and risk of PD.

#### 24. * Main outcome(s)

Give the pre-specified main (most important) outcomes of the review, including details of how the outcome is defined and measured and when these measurement are made, if these are part of the review inclusion criteria.

Our primary outcome is to compute the pooled risk of PD among beta-agonist and/or antagonist users. Studies reporting the risk ratio (RR), odds ratio (OR) and hazard ratio (HR) or the absolute numbers to estimate these measures will be of interest. Studies that qualify all other inclusion criteria and assessed any of these outcomes but did not report the data or reported the data in a format not suitable for quantitative synthesis, we will include such studies in the review and present relevant information in the narrative form

##### Measures of effect

Please specify the effect measure(s) for you main outcome(s) e.g. relative risks, odds ratios, risk difference, and/or ‘number needed to treat.

#### 25. * Additional outcome(s)

List the pre-specified additional outcomes of the review, with a similar level of detail to that required for main outcomes. Where there are no additional outcomes please state ‘None’ or ‘Not applicable’ as appropriate to the review

Not applicable

##### Measures of effect

Please specify the effect measure(s) for you additional outcome(s) e.g. relative risks, odds ratios, risk difference, and/or ‘number needed to treat.

#### 26. * Data extraction (selection and coding)

Describe how studies will be selected for inclusion. State what data will be extracted or obtained. State how this will be done and recorded.

A standard data extraction form will be adapted and used for data extraction (Appendix 3). Two reviewers will extract data independently from the included studies for the following information: study design, characteristics of the population (age, sex, and male percentage), sample size, intervention details, duration of follow-up, outcome measurements, The missing data in the literature will be obtained by emailing the corresponding author; otherwise, it will be estimated by the appropriate method according to the Cochrane Handbook 5.1.0

#### 27. * Risk of bias (quality) assessment

State which characteristics of the studies will be assessed and/or any formal risk of bias/quality assessment tools that will be used.

The methodological quality of the selected studies will be evaluated using the Jonna Briggs Institute (JBI) critical appraisal tool. A Risk-of-Bias form was adapted and will be used for data extraction. Each article will be evaluated independently by two researchers using the JBI tool. Depending on the response, each domain of the tool will be classified as high, low or unclear risk of bias.

#### 28. * Strategy for data synthesis

Describe the methods you plan to use to synthesise data. This **must not be generic text** but should be **specific to your review** and describe how the proposed approach will be applied to your data. If meta-analysis is planned, describe the models to be used, methods to explore statistical heterogeneity, and software package to be used.

The primary outcome of this study is to compute the pooled relative risk (RR) of PD among beta-agonist and/or antagonist users, separately. A generic inverse variance method will be applied to compute the overall RR of PD among beta-agonist and antagonist users. Considering the incidence of PD to be rare or low, the risk ratio, OR, and HR will be considered an equivalent measure of risk; therefore, we will use RR representing all of these measures for simplicity.14 Heterogeneity among the included studies will be assessed using Cochrane ?^2^ and I^2^ tests. Cochrane ?^2^ value (p 0.10) and I^2^ value ?50% may represent considerable heterogeneity.15 Based on the heterogeneity assessment, a random-effect or fixed-effect model will be chosen. If considerable heterogeneity observed, then the analysis will be performed using a random-effect model. Subgroup analysis will be performed based on study design, types of beta-agonist or antagonist exposure, and duration of exposure. Sensitivity analysis will be carried out by omitting a single study one by one (leave-one-out method) from the pooled analysis, and the risk of bias on the outcome will also be explored using the sensitivity analysis accordingly to the high, medium, or low risk.16 Publication bias will be detected based on the visual inspection of the funnel plot, and the trim-and-fill method will be used to estimate the effect of publication bias (if any). Meta-analysis will be performed using Review Manager version 5.4.1. (Copenhagen: The Nordic Cochrane Centre, The Cochrane Collaboration, 2014), STATA version 16 (STATA Corp., Texas, USA), and ProMeta Version 3.0 ([Computer software]. Cesena, Italy: Internovi).

#### 29. * Analysis of subgroups or subsets

State any planned investigation of ‘subgroups’. Be clear and specific about which type of study or participant will be included in each group or covariate investigated. State the planned analytic approach.

Subgroup analysis will be performed based on study design, types of beta-agonist or antagonist exposure, and duration of exposure.

#### 30. * Type and method of review

Select the type of review, review method and health area from the lists below.

##### Type of review

Cost effectiveness

No

Diagnostic

No

Epidemiologic

No

Individual patient data (IPD) meta-analysis

No

Intervention

No

Living systematic review

No

Meta-analysis

Yes

Methodology

No

Narrative synthesis

No

Network meta-analysis

No

Pre-clinical

No

Prevention

No

Prognostic

No

Prospective meta-analysis (PMA)

No

Review of reviews

No

Service delivery

No

Synthesis of qualitative studies

No

Systematic review

Yes

Other

No

##### Health area of the review

Alcohol/substance misuse/abuse

No

Blood and immune system

No

Cancer

No

Cardiovascular

No

Care of the elderly

No

Child health

No

Complementary therapies

No

COVID-19

No

Crime and justice

No

Dental

No

Digestive system

No

Ear, nose and throat

No

Education

No

Endocrine and metabolic disorders

No

Eye disorders

No

General interest

No

Genetics

No

Health inequalities/health equity

No

Infections and infestations

No

International development

No

Mental health and behavioural conditions

No

Musculoskeletal

No

Neurological

Yes

Nursing

No

Obstetrics and gynaecology

No

Oral health

No

Palliative care

No

Perioperative care

No

Physiotherapy

No

Pregnancy and childbirth

No

Public health (including social determinants of health)

No

Rehabilitation

No

Respiratory disorders

No

Service delivery

No

Skin disorders

No

Social care

No

Surgery

No

Tropical Medicine

No

Urological

No

Wounds, injuries and accidents

No

Violence and abuse

No

#### 31. Language

Select each language individually to add it to the list below, use the bin icon to remove any added in error. English

There is not an English language summary

#### 32. * Country

Select the country in which the review is being carried out. For multi-national collaborations select all the countries involved.

Australia

Czech Republic

India

#### 33. Other registration details

Name any other organisation where the systematic review title or protocol is registered (e.g. Campbell, or The Joanna Briggs Institute) together with any unique identification number assigned by them. If extracted data will be stored and made available through a repository such as the Systematic Review Data Repository (SRDR), details and a link should be included here. If none, leave blank.

#### 34. Reference and/or URL for published protocol

If the protocol for this review is published provide details (authors, title and journal details, preferably in Vancouver format)

Add web link to the published protocol.

Or, upload your published protocol here in pdf format. Note that the upload will be publicly accessible.

No I do not make this file publicly available until the review is complete

Please note that the information required in the PROSPERO registration form must be completed in full even if access to a protocol is given.

#### 35. Dissemination plans

Do you intend to publish the review on completion?

Yes

Give brief details of plans for communicating review findings.?

This systematic review meta-analysis will be published in a peer-reviewed journal.

#### 36. Keywords

Give words or phrases that best describe the review. Separate keywords with a semicolon or new line. Keywords help PROSPERO users find your review (keywords do not appear in the public record but are included in searches). Be as specific and precise as possible. Avoid acronyms and abbreviations unless these are in wide use.

Beta-agonists; beta-antagonists; beta-blockers; Parkinson’s disease

#### 37. Details of any existing review of the same topic by the same authors

If you are registering an update of an existing review give details of the earlier versions and include a full bibliographic reference, if available.

#### 38. * Current review status

Update review status when the review is completed and when it is published.New registrations must be ongoing so this field is not editable for initial submission.

Please provide anticipated publication date

Review_Ongoing

#### 39. Any additional information

Provide any other information relevant to the registration of this review.

#### 40. Details of final report/publication(s) or preprints if available

Leave empty until publication details are available OR you have a link to a preprint (NOTE: this field is not editable for initial submission). List authors, title and journal details preferably in Vancouver format.

Give the link to the published review or preprint.

## References

1. de Germay S, Conte C, Rascol O, Montastruc J-L, Lapeyre-Mestre M. β-Adrenoceptor Drugs and Parkinson’s Disease: A Nationwide Nested Case–Control Study. CNS drugs 2020;34:763–72.

2. Jankovic J, Tan EK. Parkinson’s disease: etiopathogenesis and treatment. Journal of Neurology, Neurosurgery & Psychiatry 2020;91:795–808.

3. Kouli A, Torsney KM, Kuan W-L. Parkinson’s disease: etiology, neuropathology, and pathogenesis. Exon Publications 2018:3–26.

4. N. Maserejian, L. Vinikoor-Imler, A. Dilley. Estimation of the 2020 Global Population of Parkinson’s Disease (PD) [abstract]. Mov Disord. 2020; 35 (suppl 1). https://www.mdsabstracts.org/abstract/estimation-of-the-2020-global-population-of-parkinsons-disease-pd/. Accessed April 21, 2021.

5. Fields CR, Bengoa-Vergniory N, Wade-Martins R. Targeting alpha-synuclein as a therapy for Parkinson’s disease. Frontiers in molecular neuroscience 2019;12:299.

6. Mittal S, Bjørnevik K, Im DS, et al. β2-Adrenoreceptor is a regulator of the α-synuclein gene driving risk of Parkinson’s disease. Science 2017;357:891–8.

7. Chen W, Sadatsafavi M, Tavakoli H, Samii A, Etminan M. Effects of β2-Adrenergic Agonists on Risk of Parkinson’s Disease in COPD: A Population-Based Study. Pharmacotherapy: The Journal of Human Pharmacology and Drug Therapy 2020;40:408–15.

8. Hopfner F, Wod M, Höglinger GU, et al. Use of β2-adrenoreceptor agonist and antagonist drugs and risk of Parkinson disease. Neurology 2019;93:e135–e42.

9. Koren G, Norton G, Radinsky K, Shalev V. Chronic Use of β-Blockers and the Risk of Parkinson’s Disease. Clinical drug investigation 2019;39:463–8.

10. Page MJ, McKenzie JE, Bossuyt PM, et al. The PRISMA 2020 statement: an updated guideline for reporting systematic reviews. BMJ 2021;372:71.

11. Stroup DF, Berlin JA, Morton SC, et al. Meta-analysis of observational studies in epidemiology: a proposal for reporting. Meta-analysis Of Observational Studies in Epidemiology (MOOSE) group. Jama 2000;283:2008–12.

12. Hussain S, Singh A, Baxi H, Taylor B, Burgess J, Antony B. Thiazolidinedione use is associated with reduced risk of Parkinson’s disease in patients with diabetes: a meta-analysis of real-world evidence. Neurological sciences 2020;41:3697–703.

13. Cumpston M, Li T, Page MJ, et al. Updated guidance for trusted systematic reviews: a new edition of the Cochrane Handbook for Systematic Reviews of Interventions. Cochrane Database Syst Rev 2019;10:ED000142.

14. Knott C, Bell S, Britton A. Alcohol Consumption and the Risk of Type 2 Diabetes: A Systematic Review and Dose-Response Meta-analysis of More Than 1.9 Million Individuals From 38 Observational Studies. Diabetes care 2015;38:1804–12.

15. Singh A, Hussain S, Najmi AK. Number of studies, heterogeneity, generalisability, and the choice of method for meta-analysis. Journal of the neurological sciences 2017;381:347-.

16. Krzanowski W, Hand D. Assessing error rate estimators: the leave-one-out method reconsidered. Australian Journal of Statistics 1997;39:35–46.

17. Guyatt G, Oxman A, Vist G, Kunz R, Falck-Ytter Y, AlonsoCoello P, et al. GRADE: an emerging consensus on rating quality of evidence and strength of recommendations. BMJ 2008;336(7650):924–6.

